# A linked mixture model of coronary atherosclerosis

**DOI:** 10.1101/2021.09.13.21263547

**Authors:** Bret Beheim

## Abstract

**Background & Objectives:** Characterizing the progression of coronary atherosclerosis is a critical public health goal. The most common quantitative summary, the CAC score, is modelled by a variety of statistical methods, both as a predictor of coronary events and as an outcome of behavioral and population-specific risk factors. Little attempt has been made, however, to ground these statistical models in the underlying physiology of arterial aging, which would allow us to describe the onset and growth of CAC over a patient's life.

**Methods:** Using a generative growth model for arterial plaque accumulation, we identify severe under-estimation in the age of initial onset and rate of progression (doubling time) of CAC growth with standard ln(CAC + 1) or ln(CAC | CAC > 0) models, and use this growth model to motivate new statistical approaches to CAC using logistic and log-linear mixture regressions. We compare statistical models directly by computing their average parameter biases using 540 growth trajectory simulations (113,760 patients, 268,200 observations).

**Results:** While all models used can successfully estimate the influence of risk factors with minimal bias, we demonstrate substantial improvements in predictive accuracy in the timing of CAC onset and progression with logistic regression and linked hurdle-lognormal mixture regression, compared with standard ln(CAC + 1) or ln(CAC | CAC > 0) models.

**Conclusions:** Using models that can account for patient-specific onset and progression rates, accurate descriptions of CAC trajectories can be made even in cross-sectional (single scan per patient) designs, with substantial clinical and epidemiological utility.

## I. INTRODUCTION

The coronary calcium (CAC) score was first proposed by Agatston, *et al*. [1] to quantify the amount of calcified plaque in the coronary arteries, and remains the standard metric for assessing arterial aging [2, 3]. Three decades of research with large cohorts of patients, in both cross-sectional and longitudinal designs, has shown a patient’s CAC score to be highly predictive of subsequent major coronary events [4–8], even among patients with few additional risk factors [9, 10]. Many studies have found CAC varies systematically by nationality [11, 12] and ethnicity [13–16], with some populuations showing very little CAC even into advanced age [17], leading to debates over whether atherosclerosis itself is a consequence of sedentary, industrialized lifestyles [18, 19].

Despite the centrality of the CAC score in the study of atherosclerosis, its significance in clinical practice is ambiguous [20, 21], and it lacks a consistent statistical model from study to study. A given analysis may employ ordered-categorical [8, 22–24], linear [25, 26], log-linear [21, 27–29], quantile [30, 31] and proportional hazards [32–34] regression models. With some exceptions, e.g. [26, 30, 35–38], the CAC score is almost always modelled as a cross-sectional value that varies between patients, rather than as an atherosclerotic growth process within an individual.

To better connect the CAC score to the dynamic physiology of arterial aging, we propose a simple growth model for the onset and progression of calcified arterial plaques. Using high-throughput computational simulations to generate 540 realistic CAC studies (360 cross-sectional and 180 longitudinal studies, 113,760 patients, 268,200 observations), this model can identify structural mis-specifications in many of the common log-linear models of CAC, and indicate specific statistical modifications which dramatically improve the predictive accuracy of CAC modelling across the life course.

## II. METHODS

### A generative model of CAC growth

Although CAC is modelled in the current literature described above by a large variety of methods, we can synthesize two key empirical details from which to build a generative model [39]. First, calcified plaques typically appear in the coronary artery in middle age, and before a patient’s age of onset, they will show a CAC score of 0 if given a computerized tomography scan. After the age of onset, an individual’s CAC steadily accumulates at one or more plaque sites in the coronary artery, increasing at a non-linear pace set in part by variation in genetics, behavior, or risk exposures [9, 26, 35, 40], with CAC measurements of longitudinal study participants increasing between 14.3% and 29.7% annually [41, 42]. Given this, let us assume that the rate of growth in CAC at a given moment *t* after the age of onset *t*_0_ is proportional to both immediate risk factors and the current amount of coronary calcium, that is,

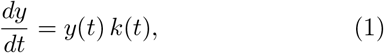

for CAC score *y*(*t*) and positive growth function *k*(*t*). The CAC score is thus given by the integral of all growth velocities since the age of onset *t*_0_,

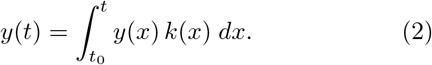

In principle, we could estimate the shape of the rate function *k*(*t*) as we observe the score *y* change with time. However, since we ordinarily do not have more than one CAC measurement per patient, we can instead appeal to the *average* growth rate between *t*_0_ and *t*,

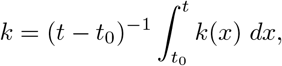

a constant. We can then solve Equation 1 as

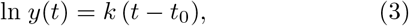

for any *t* ≥ *t*_0_. Since rate *k* describes an exponential growth process, it is useful to re-express *k* as a *doubling time*. That is, for any CAC score *y* at time *t*, we can describe the time *t*+*d* at which the score is doubled, which is true if *d* = (ln 2)*/k*. Longitudinal studies of CAC report an average doubling time between 2.7 and 5.8 years [41, 42]. An example atherosclerosis trajectory and linear approximation following this approach for a single patient is given in Figure 1.

**FIG. 1.**
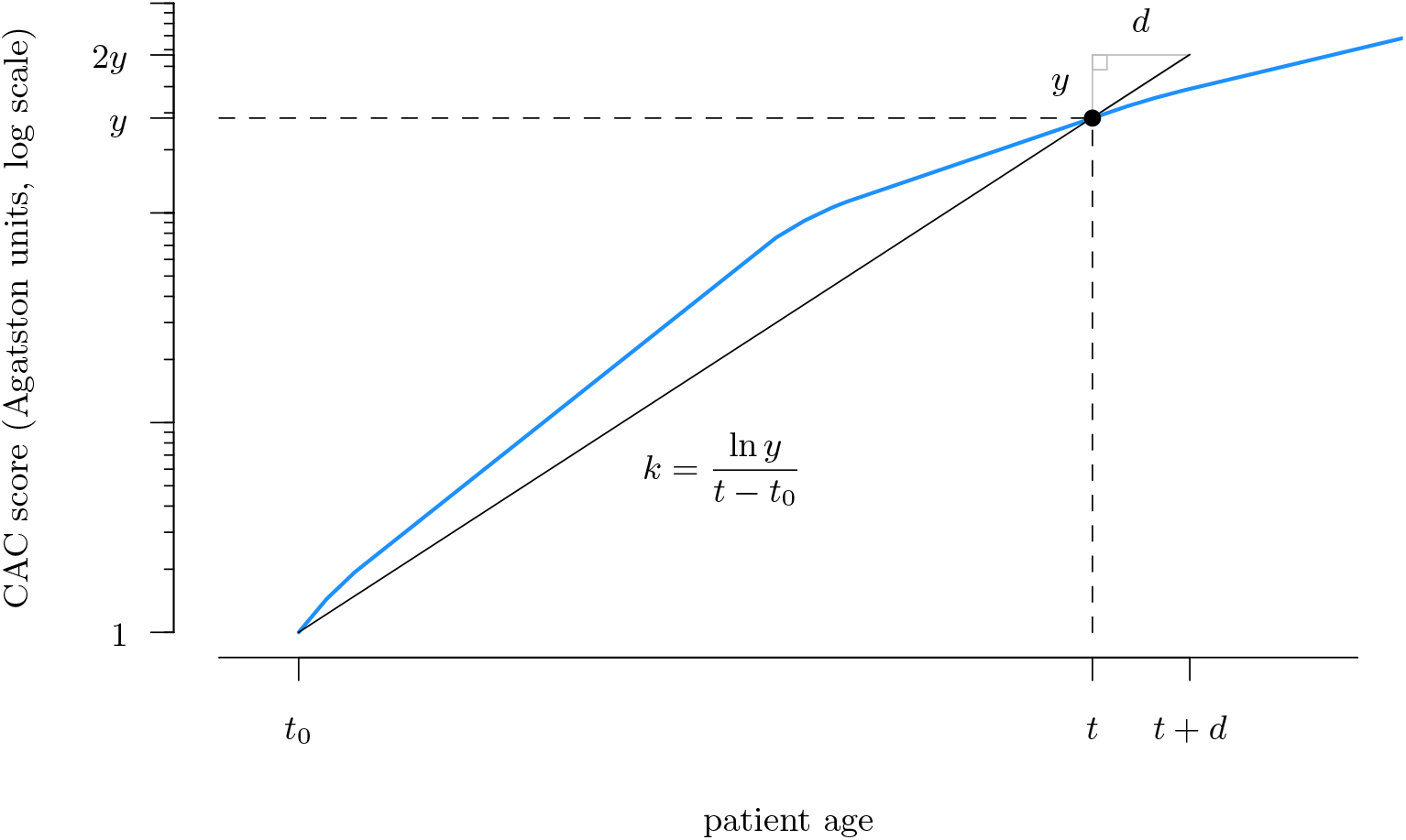
A single patient’s coronary atherosclerosis progression over time, as measured by CAC score (blue). Onset at age *t*_0_ is defined as the initial appearance of calcified plaque (CAC>0), which is equivalent to a CAC score of 1 in most studies. Before *t*_0_ the patient’s CAC is 0. All CAC measurements after *t*_0_ are given by Equation 2. Average growth rate *k* and doubling time *d* = ln(2)*/k* are defined by a linear approximation (black line) using the location of *t*_0_ and observation *y*. In this growth trajectory, the patient’s instantaneous CAC growth rate *k*(*t*) intermittently decreases. Because we are usually confined to a single measurement *y* at some time *t*, neither the location of *t*_0_ nor the rate function *k*(*t*) are directly observable, but can be approximated by log-linear regression methods (Equations 4, 5).

Allowing for a small amount of measurement error, we can estimate the average *k* for a set of observations by log-linear regression. For each observation *j*, with CAC score *y*_*j*_, and patient age *t*_*j*_,

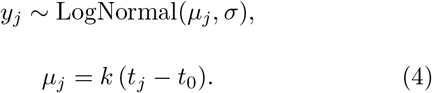

That is, each observation varies around some average CAC score *µ*_*j*_ with standard deviation *σ* due to measurement error and other unobserved factors. Given a dataset recording a single individual’s CAC trajectory over time, and the log-linear regression model *µ*_*j*_ = *a*+*bt*_*j*_, we could estimate the model parameters in Equation 4 as *t*_0_ = (−*a/b*), *k* = *b* and *d* = ln(2)*/b*.

### Structural mis-estimation of disease onset and progression

Studying CAC on the logarithmic scale was originally proposed by Agatston, *et al*. [1] and remains the most common statistical approach, commonly with ln(CAC+1) or ln(CAC|CAC>0) as dependent variables. However, the growth model described in Equation 4 indicates the existence of two major estimation biases with such methods (Figure 2). Principally, the existence of distinct CAC age trajectories across patients is not a structural component of log-linear regression, which treats each patient-observation as following a single population average log-linear path from a single age of onset, *t*_0_. As a consequence, standard regression models of CAC systematically under-estimate the age of atherosclerotic onset (Figure 2, left) and rate of progression (Figure 2, right) in direct proportion to between-patient variability. Under realistic growth conditions, a standard deviation in atherosclerotic onset among patients of only 3 years can induce estimation biases as severe as five decades.

**FIG. 2.**
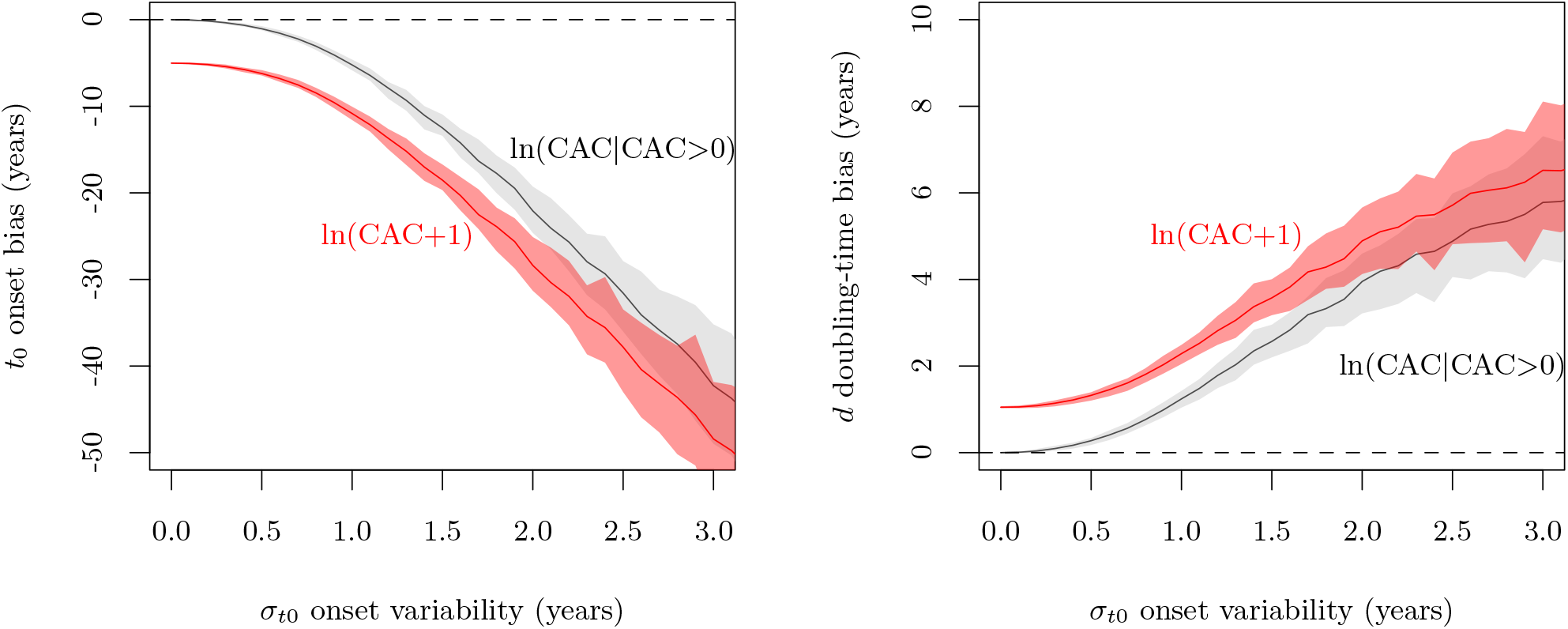
The relationship between inter-individual variance in *t*_0_ and bias in linear regression estimates of the age of onset (left) and the doubling time (right) of CAC for a simulated cohort of 5,000 patients (*t*_0_ = 40, *k* = 1). Experimentally controlling the simulation standard deviation in individual age of onset, 100 cross-sectional datasets are constructed, each selecting a single random observation from each patient growth trajectory (average scan age 60*±*10 years). For each dataset, parameter estimates are calculated in a log-linear regression of ln(CAC+1) (red) or ln(CAC|CAC>0) (black) on patient age, and the true onset age *t*_0_ in the simulation is subtracted from onset estimates −*a/b*. Average cross-sectional estimates are shown with 89% confidence regions. The ln(CAC+1) estimates are, on average, -5.8 years more biased than ln(CAC |CAC>0) on *t*_0_ and 0.9 years more biased in *d* over the observed range of *σ*_*t*0_. Reproducible R code for this simulation and figure is available in the Supplementary Materials.

An additional bias is introduced by studies that alter the scores of patients with no atherosclerosis (CAC = 0). Since 0 is undefined on the logarithmic scale, many studies first re-assign these patients a CAC score of 1 [6, 12, 16, 21, 28, 29, 43–47], indicating these patients have already begun atherosclerosis. The common justification given for the log-transform is to normalize the distribution of CAC [1, 12, 16, 33]. Yet depending on the cohort, a majority of patients may have a CAC score of 0. Consequently, such logarithmic outcome distributions are extremely zero-inflated, e.g. Figure 1 in [12]. By estimating the average growth rate *k* from a realistic population in which some are growing CAC at rate *k* = 1 and others are not growing CAC at all, we can expect loglinear regression models to introduce additional biases of at least -5.8 years in CAC age of onset (Figure 2, left, red) and 0.9 years in CAC doubling time (Figure 2, right, red) versus studies that exclude non-atherosclerotic patients before performing the regression [15, 17, 26, 27, 48, 49].

### Alternate statistical specifications

To prevent inter-individual variation from manifesting as bias in statistical analyses, we can modify the loglinear model in Equation 4 with explicit CAC growth trajectories for each patient *i* by

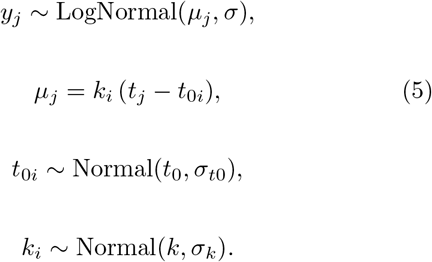

That is, the time of initial onset for each individual *i, t*_0*i*_, is assumed to vary around some population-average age *t*_0_ by standard deviation *σ*_*t*0_, and similarly for individual-specific average growth rate *k*_*i*_.

Rather than exclude patients who have no CAC, or introduce bias by altering their values, another approach is to employ generalized linear regression to model the probability CAC>0 [4, 5, 8–11, 32, 38, 50–55]. If we define the log-odds that a patient’s CAC score at observation *j* is non-zero as *θ*_*j*_ = logit(Pr(*y*_*j*_>0)), then for any individual *i* observed at age *t*_*j*_,

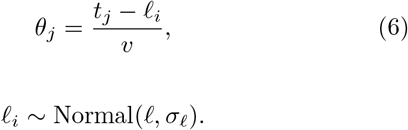

The probability CAC is observed to be nonzero rises sinusoidally from 0 to 1 over the life course, reaching 50% probability at age *ℓ*_*i*_ for patient *i*. We can expect *ℓ*_*i*_ to vary by individual according to parameters *v* and *σ* _*ℓ*_, but also systematically by risk factors, e.g. patient sex, which can be incorporated into a multivariate logistic regression model. Unlike log-linear models above, this approach can utilize all observations, including those for which *y*_*j*_ = 0, at the cost of being unable to distinguish non-zero CAC magnitudes.

Models of CAC>0 can be combined with models of CAC magnitudes in so-called mixture models [56], first introduced in the CAC literature by Kaplan, *et al*. [17] with the use of a zero-inflated Negative Binomial model. Such models incorporate two distinct probability distributions to describe both zero outcomes and non-zero magnitudes simultaneously. Here we employ the related concept of a hurdle model, which allows an additional link between these two distributions. Specifically, in modelling the probability of CAC>0 by age, we are simultaneously describing the age of onset as a random quantity. If we re-interpret the inverse-logit link as a cumulative distribution function, onset age for each individual *i* must follow a Logistic probability distribution with mean *ℓ*_*i*_ and standard deviation 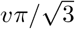. Therefore, both the hurdle and log-normal components of this mixture share a common parameter in the individual onset age, as

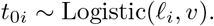

Thus, the ages of onset in a multi-trajectory log-linear model are random quantities that follow the parameters of the hurdle model. During Bayesian statistical estimation, the hurdle component’s *ℓ*_*i*_ and *v* are updated by CAC presence/absence observations, adding additional resolution on the location of *t*_0*i*_ in the log-linear model. In turn, the log-linear estimation of *t*_0*i*_ is updated by observed magnitudes of nonzero CAC, and so inform the locations of *ℓ*_*i*_ and *v* in the hurdle component (Figure 3). Taken together, this fully defines the linked hurdle-lognormal mixture model. A complete list of variables is given in Table III.

**FIG. 3.**
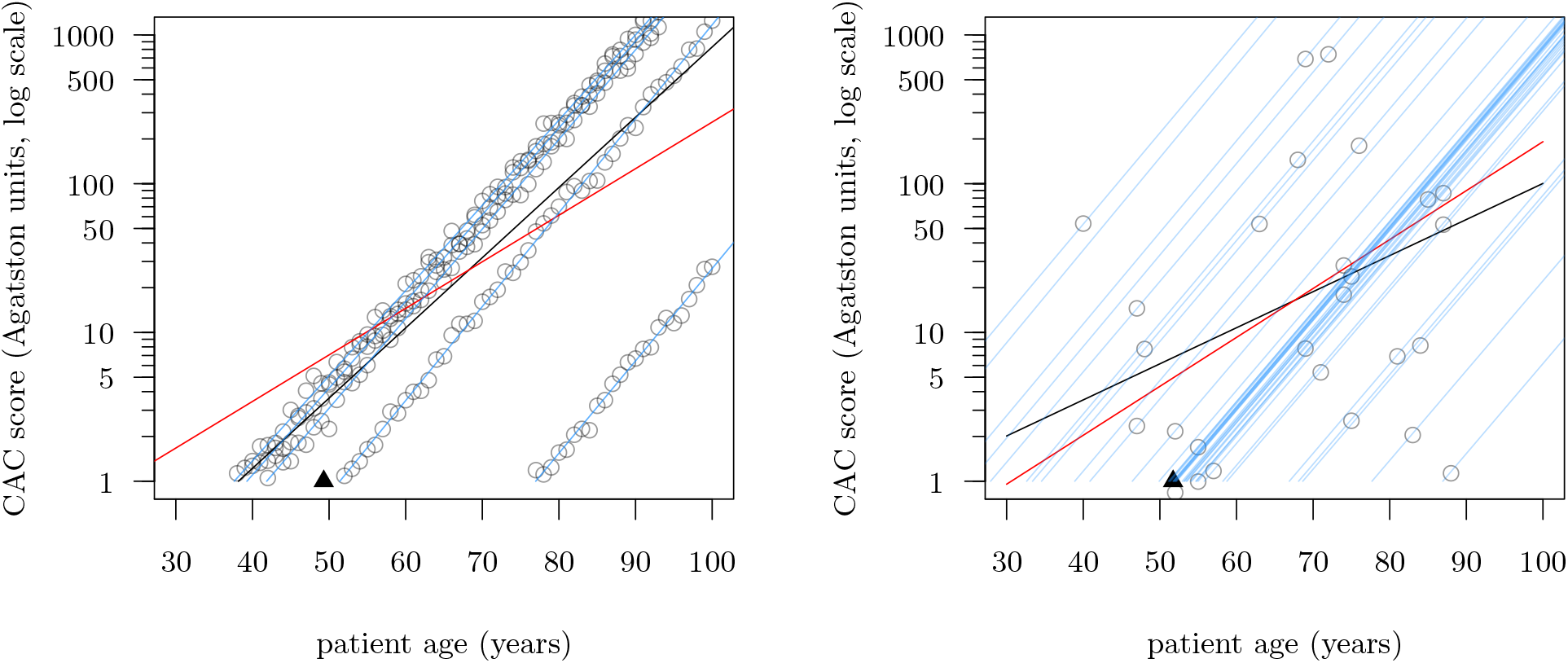
Observed CAC trajectories for 5 longitudinal study patients (left) and 40 cross-sectional patients (right), with average counterfactual predictions from fitted ln(CAC|CAC>0) (black), ln(CAC+1) (red) and linked hurdle-lognormal (blue) models. In each figure, the true population average *t*_0_ is indicated by a black triangle. All statistical models are fit in R and Stan, with prior uncertainty equal to simulation uncertainty. Reproducible R code for this simulation and figure is available in the Supplementary Materials.

### Ranking statistical models by generative simulation

In order to characterize gross performance differences between the above methods of studying CAC growth, we simulated 540 datasets for populations experiencing a variety of realistic atherosclerotic growth scenarios (Table I), varying in the number of patients observed (from 1 to 1,000 patients), the number of observations per patient and the impact of standard risk factors (age and sex) on initial onset and progression. We can summarize patient data for N = 113,760 patients seen over all 540 studies in Table II.

**TABLE I.** Tabulation of conditions across 540 simulated CAC studies. Here “longitudinal” studies involve 100 annual observations over the entire course of each patient’s life (all individuals live to age 100), while “cross-sectional” studies simulate realistic CAC datasets by randomly sampling a single observation from each patient’s adult measurements. As sex is a commonly-observed risk factor across studies, some populations here include a root-mean-square difference between men and women of 5.7 years on *t*_0_ and 1.4 years on *d*. If present, between-individual variation is set by model parameters *σ*_*t*0_ = 4.0 years and *σ*_*d*_ = 1.0 years. Bayesian statistical models use either “simulation” priors, set to have the same uncertainty as the generative simulation itself, or “regularizing” priors, which reflect larger, empirically-plausible ranges for each parameter [56]. Generative R code to create these simulated populations can be found in the Supplementary Materials.

|  | Longitudinal | Cross-sectional |
| --- | --- | --- |
| Total Studies | 180 | 360 |
| Inter-Individual Variation |  |  |
| No Variation | 90 | 0 |
| Variation on $t_0, d$ | 90 | 180 |
| Sex Effect |  |  |
| No sex differences | 60 | 120 |
| Sex difference in $t_0$ | 120 | 240 |
| Sex difference in $t_0, d$ | 60 | 120 |
| Prior Accuracy |  |  |
| Regularizing Priors | 90 | 180 |
| Simulation Priors | 90 | 180 |
| Study Size |  |  |
| $N = 1$ Patient | 60 | 0 |
| $N = 5$ Patients | 60 | 0 |
| $N = 20$ Patients | 60 | 60 |
| $N = 50$ Patients | 0 | 60 |
| $N = 100$ Patients | 0 | 60 |
| $N = 200$ Patients | 0 | 60 |
| $N = 500$ Patients | 0 | 60 |
| $N = 1000$ Patients | 0 | 60 |

**TABLE II.** Characteristics of patients across 360 cross-sectional and 180 longitudinal CAC studies. Means (standard deviations) are given for *t*_0_ and *d*.

|  | Women | Men | Total |
| --- | --- | --- | --- |
| Number of Patients | 56,901 | 56,859 | 113,760 |
| Longitudinal | 786 | 774 | 1,560 |
| Cross-sectional | 56,115 | 56,085 | 112,200 |
| Number of Observations | 134,715 | 133,485 | 268,200 |
| Longitudinal | 78,600 | 77,400 | 156,000 |
| Cross-sectional | 56,115 | 56,085 | 112,200 |
| Onset $t_0$ (years) | 49.8 (10.3) | 50.1 (10.4) | 49.9 (10.3) |
| Progression $k$ | 1.5 (0.3) | 1.5 (0.3) | 1.5 (0.3) |
| Doubling time $d$ (years) | 5.0 (1.1) | 5.0 (1.1) | 5.0 (1.1) |
| CAC category (%) |  |  |  |
| 0 | 42.8 | 43.0 | 42.9 |
| 1 to 9 | 20.4 | 20.6 | 20.5 |
| 10 to 99 | 19.0 | 19.1 | 19.1 |
| 100 to 399 | 8.1 | 8.0 | 8.1 |
| 400 to 999 | 3.6 | 3.5 | 3.5 |
| $\geq 1000$ | 5.7 | 5.4 | 5.6 |

**TABLE III.** Glossary of variables used.

| Variable | Description |
| --- | --- |
| $k(t)$ | instantaneous growth rate in CAC at age $t$ |
| $y(t)$ | true patient CAC score at age $t$ |
| $y_j$ | observed CAC score at observation $j$ |
| $t_j$ | patient age at observation $j$ |
| $t_0$ | population average age of onset |
| $t_{0i}$ | age of onset for patient $i$ |
| $k$ | population average growth rate in CAC |
| $d$ | population average doubling time in CAC |
| $k_i$ | average growth rate in CAC for patient $i$ |
| $d_i$ | average doubling time in CAC for patient $i$ |
| $a$ | log-linear regression intercept |
| $b$ | log-linear regression slope |
| $v$ | scaling term for log-odds of CAC $> 0$ |
| $\theta_j$ | log-odds of CAC $> 0$ for patient $i$ in measurement $j$ |
| $\ell$ | expected population average age of onset |
| $\ell_i$ | expected age of onset for patient $i$ |
| $\sigma$ | standard deviation on each CAC measurement $y_j$ |
| $\sigma_{t0}$ | standard deviation on $t_{0i}$ |
| $\sigma_k$ | standard deviation on $k_i$ |
| $\sigma_\ell$ | standard deviation on $\ell_i$ |

For each study, we consider each of the seven models described above: log-linear regressions of ln(CAC+1) or ln(CAC |CAC>0) on patient age and logistic regressions of Pr(CAC>0) with or without individual trajectories for each patient *i*, and mixture models including a logistic hurdle and log-linear magnitude component with or without a hierarchical link between *t*_0*i*_ and *ℓ*_0*i*_. The seven models were fit to each of the 540 datasets, statistical estimates were calculated and extracted, and, given the true values for each individual trajectory were known, the average bias of each parameter estimate was computed. To assess bias in estimating the importance of other risk factors besides patient age, we include patient sex (male or female) as an additional covariate in each model. All simulations were performed in R version 4.0.4 and models were fit with Stan by the cmdstanr library version 0.3.0.9, with all analysis code available in the Supplementary Materials.

## III. RESULTS

Model performance is summarized in Table IV. Traditional log-linear models of ln(CAC+1) and ln(CAC|CAC>0) show large, systematic estimation errors across cross-sectional studies, of -11.5 years (SE: 0.2, P*<*0.001) and -9.7 years (SE: 0.3, P*<*0.001), respectively. Similarly, these models systematically under-estimate the average rate of progression, with a doubling-time bias of 2.4 years (SE: 0.1, P*<*0.001) for ln(CAC+1) and 1.8 years (SE: 0.1, P*<*0.001) for ln(CAC|CAC>0) over the true doubling time.

**TABLE IV.**
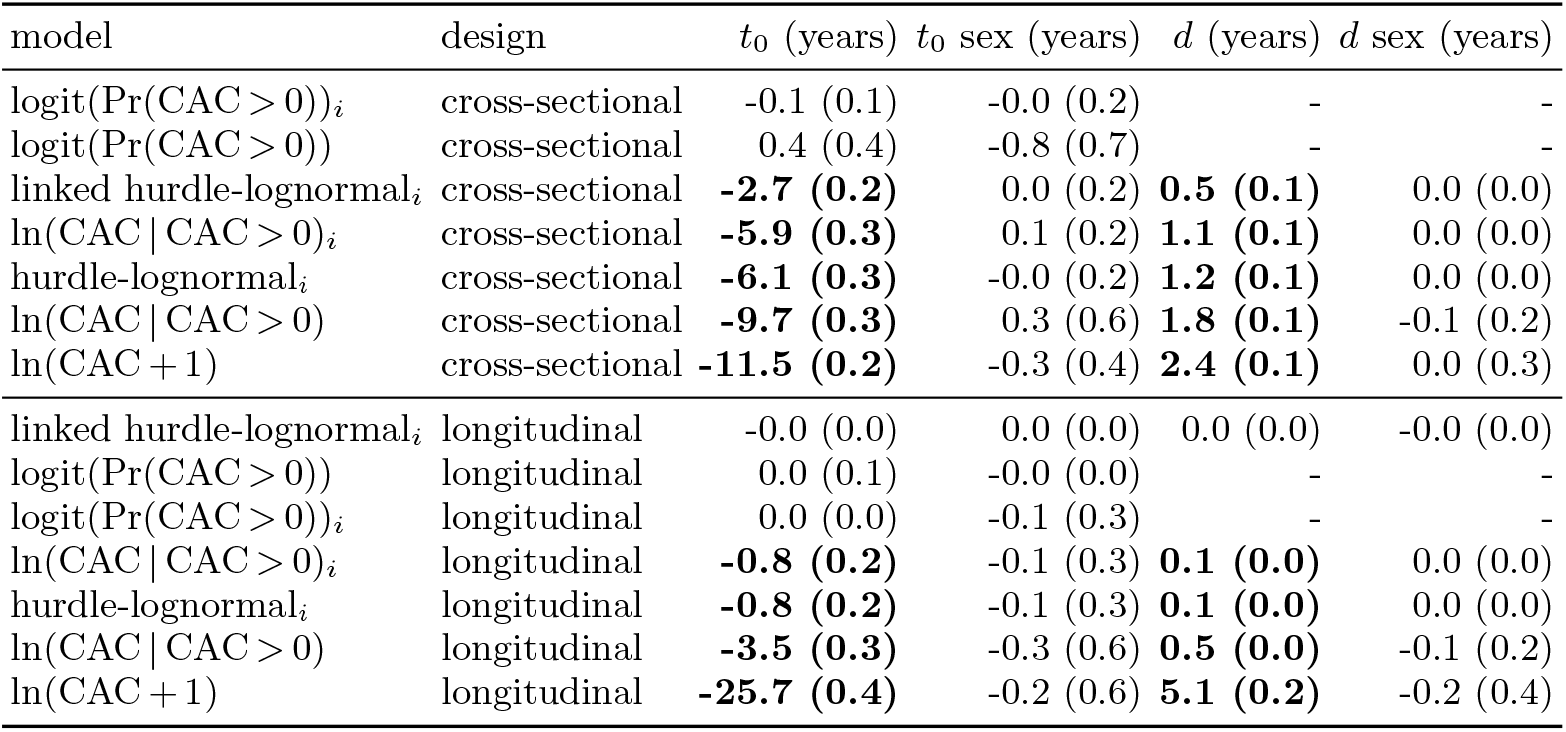
Estimation bias (mean and standard error) over all 540 studies, in years. Statistical models that include individual-specific trajectories (subscript *i*) calculate bias estimates using the true *t*_0*i*_ and *d*_*i*_ values of each patient; models without individual-specific trajectories estimate population-level parameters, which are compared against the true population average *t*_0_ and *d* within each simulated study. Biases on “sex” represent the estimated average difference between men and women minus the true generative difference (or lack thereof). Since E(*t*_0*i*_) = *ℓ*_*i*_, logistic regression models use *ℓ*_*i*_ or *ℓ* as equivalents to *t*_0*i*_ and *t*_0_. Estimates in boldface are statistically distinct from zero bias by two-tailed *t*-tests with *α* = 0.001.

In both cross-sectional and longitudinal designs, estimation of onset ages (population average *t*_0_ and patient-specific *t*_0*i*_) is least biased in models that account for CAC presence / absence. In cross-sectional designs, a logistic regression model estimating the population average age of onset *t*_0_ is biased by 0.4 years (SE: 0.4), and not statistically distinct from zero bias (P=0.367). Logistic regression estimates of individual patient ages *t*_0*i*_ are similarly biased by only -0.1 years (SE: 0.1, P=0.430). The linked hurdle log-normal model is biased in cross-sectional designs on average -2.7 years (SE: 0.2, P*<*0.001) but out-performs all other log-linear models in estimation of both age of onset and in estimation of the doubling time.

Less systematic bias is observed across models in the estimation of the importance of sex as a predictor of onset and progression. In both cross-sectional and longitudinal designs, all seven models were able to assess the true covariate effect without clear bias, although standard loglinear models show the largest standard errors in bias in both *t*_0_ and *d* (Table IV).

## IV. DISCUSSION

Generally speaking, logistic regression and mixture models out-perform log-linear models in estimating population and individual-specific ages of CAC onset, and models that explicitly account for individual patient trajectories out-perform those that do not. These advantages are most pronounced in cross-sectional study designs, which constitute the vast majority of existing CAC datasets.

Because logistic regression models are able to accurately utilize the full set of patient observations, including those from the period before CAC growth begins, they are able to characterize the onset of atherosclerosis more accurately than log-linear models, which must either exclude patients observed with zero CAC or incorrectly alter these scores to accomodate the logarithmic scale, producing large estimation errors.

Although large biases in estimation are observed in some models in patient-specific CAC onset and progression, substantially smaller, unsystematic errors are found in estimation of the impact of sex. As a proxy for other risk factors such as smoking, obesity and physical activity, this covariate’s predictive value is thus less likely to be altered significantly by choice of model in realistic datasets.

While the innovations in model design described above show marked improvements in estimation accuracy, they come with increased costs, in terms of model complexity and computation time, and successful posterior updating of the full linked-hurdle log-normal may not be feasible for very large samples of patients. However, Table IV indicates that estimation performance can be improved for specific parameters without the use of the full linked hurdle-lognormal model: for parameter *d*, the ln(CAC|CAC>0)_*i*_ model is, on average, less biased than either standard log-linear model, and for *t*_0_, the logistic regression models with or without individual trajectories are both very accurate.

More generally, although the simulation studies presented here cover a broad range of realistic epidemiological scenarios, they are strictly limited to the context of the generative model described by Equations 1 and 2. The true process of CAC onset and progression is necessarily more complex, and is only part of the larger process of coronary atherosclerosis, which can be seen as beginning with arterial lesions, and progress to stenosis and vascular occlusion even in the absence of CAC [3, 57– 59]. Incorporating more physiological details, such as the process of soft plaque accumulation prior to calcification, plaque density and stability [60] or the site-specific formation of individual calcified plaques [20], can potentially lead to further modelling innovations, and consequently greater improvements in estimation accuracy.

## V. CONCLUSION

Currently, the study of atherosclerosis involves a wide variety methods of modelling the CAC score, with multiple statistical designs employed even within the same study. Both because of this awkwardness, and because the CAC score only suimmarizes the underlying distribution of calcified plaque, some have suggested alternative metrics to studying atherosclerosis progression [20, 21]. However, the above computational meta-analysis shows the current statistical approaches to CAC modelling have substantial structural biases, tending to represent atherosclerotic growth as beginning earlier and progressing slower than it really does. Modifications to these models that recognize patient-specific growth trajectories are substantially more accurate in realistic simulation studies with both longitudinal and cross-sectional designs, and so we can expect such models to similarly out-perform traditional methods in the analysis of real patient data.

## Data Availability

All statistical models and simulation data and software are available in the following github repository.

https://github.com/babeheim/linked-calcium-growth

## Acknowledgments

Hillard Kaplan, Michael Gurven, Gregory Thomas, Michael Miyamoto, Richard McElreath, Cody Ross, Randall C. Thompson, Margaret Gatz, L. Samuel Wann, Adel H. Allam & Andrei Irimia provided valuable feedback on earlier versions of this analysis.

## Supplementary materials

All statistical models and simulation data and software are available at https://github.com/babeheim/linked-calcium-growth.

